# Higher viral load drives infrequent SARS-CoV-2 transmission between asymptomatic residence hall roommates

**DOI:** 10.1101/2021.03.09.21253147

**Authors:** Kristen K. Bjorkman, Tassa K. Saldi, Erika Lasda, Leisha Conners Bauer, Jennifer Kovarik, Patrick K. Gonzales, Morgan R. Fink, Kimngan L. Tat, Cole R. Hager, Jack C. Davis, Christopher D. Ozeroff, Gloria R. Brisson, Daniel B. Larremore, Leslie A. Leinwand, Matthew B. McQueen, Roy Parker

## Abstract

In 2019-2020, the COVID-19 pandemic spread to over 200 countries in less than six months. To understand the basis of this aggressive spread, it is essential to determine the transmission rate and define the factors that increase the risk of transmission. One complication is the large fraction of asymptomatic cases, particularly in young populations: these individuals have viral loads indistinguishable from symptomatic people and do transmit the SARS-CoV-2 virus, but they often go undetected. As university students living in residence halls commonly share a small living space with roommates, some schools established regular, high density testing programs to mitigate on-campus spread. In this study, we analyzed longitudinal testing data of residence hall students at the University of Colorado Boulder. We observed that students in single rooms were infected at a lower rate than students in multiple occupancy rooms. However, this was not due to high rates of transmission between roommates, which only occurred approximately 20% of the time. Since these cases were usually asymptomatic at the time of diagnosis, this provides further evidence for asymptomatic transmission. Notably, individuals who likely transmitted to their roommates had an average viral load ∼6.5 times higher than individuals who did not. Although students were moved to separate isolation rooms after diagnosis, there was no difference in time to isolation between these cases with or without transmission. This analysis argues that inter-roommate transmission occurs in a minority of cases in university residence halls and provides strong correlative evidence that viral load can be proportional to the probability of transmission.

## INTRODUCTION

As of February 28, 2021, over 2.5 million people worldwide had died from Coronavirus Disease-19 (COVID-19), the disease caused by the novel coronavirus SARS-CoV-2 (CSSE at Johns Hopkins University, 2021; Dong et al., 2020). As an airborne respiratory disease, households are at elevated risk due to close personal contact and a shared living environment. However, secondary attack rates are estimated at 17%, which indicates that many households do not experience spread (Madewell et al., 2020). The factors underlying this variable transmission risk are a matter of intense scrutiny. Although many studies have found a broad range in viral load over 6 orders of magnitude or more (Kissler et al., 2020; Lennon et al., 2020; Yang et al., 2021), whether this contributes to the risk of transmission is unclear. In addition, time spent with a close contact is also a potential contributing factor, but understanding its impact is hampered by differences in isolation practices and variability in the timing of case detection.

The high prevalence of asymptomatic infections can be a confounding variable as many studies focus primarily on symptomatic index cases, particularly those that have been hospitalized. This is most likely because asymptomatic cases are frequently under-reported due to lack of overt illness revealing them and inadequate testing resources to look for them. However, studies with high testing penetrance of large populations indicate that 40-90% of cases may be asymptomatic (Lavezzo et al., 2020; Lennon et al., 2020; Poletti et al., 2020). Importantly, asymptomatic infected individuals still transmit virus and show no difference in peak viral load compared to symptomatic individuals, and they are also expected to be more mobile because they are not experiencing illness (Cevik et al., 2021; Lavezzo et al., 2020; Lee et al., 2020; Yang et al., 2021). Therefore, it is critical to study this potentially large reservoir of viral carriers.

Roommates living in college and university residence halls are an example of households with close contacts: they typically share a bedroom, bathroom, and dining facilities. Moreover, they are also a young adult population with a higher likelihood of being asymptomatic and a lower rate of co-morbidities that would elevate risk of severe disease (Lennon et al., 2020; Poletti et al., 2020; Williamson et al., 2020). Thus, they present an ideal opportunity to analyze the extent of transmissibility from asymptomatic or mildly symptomatic cases.

During the Fall 2020 academic semester (August 17 – November 25), the University of Colorado Boulder (CU Boulder) followed a traditional model of on-campus living, including roommate pairing. This was accompanied by a student-centered public health system requiring mandatory weekly RT-qPCR screening of all asymptomatic residential students, thus providing a record of infection events within the residence halls while also identifying students who remained uninfected due to continued negative testing. Infected students identified through RT-qPCR screening were referred for confirmatory diagnostic RT-qPCR testing at the student health center. Additional diagnostic testing of students in the residence halls occurred when students exhibited symptoms or were identified through contact tracing as being exposed to COVID-19. Students living in the residence halls with confirmed cases of COVID-19 were either housed in on-campus isolation or returned to their permanent off-campus residence to isolate for 10 days.

Aggressive screening with a quantitative molecular assay and a robust track/trace/isolate system allowed us to examine the factors that may influence transmissibility in a household, particularly from asymptomatic individuals. Therefore, we determined (1) the extent of roommate transmission, (2) the contribution of viral load to the likelihood of spread, and (3) the impact of time spent in the room while infected before moving to temporary isolation.

## RESULTS

At residence hall move-in between August 17 and August 21, 2020, residential students at CU Boulder were required to either provide proof of recent negative RT-qPCR test (no earlier than 5 days prior) or participate in on-campus RT-qPCR testing on the day of their arrival. This was an effort to minimize infected students moving into the residence halls. Thereafter, students were required to test at least weekly but were exempt from this mandate for the rest of the semester after a COVID-19 diagnosis. The first cases in residence halls were detected by August 24, and 1058 positive residential students were ultimately detected by November 25, after which time residence halls were closed as scheduled for winter break. This represented 16.5% of the 6408 residential students. Many cases in the residence halls originated through off-campus contacts based on case investigation and contact tracing (CU Pandemic Response Office, manuscript in preparation), but cases in residence halls had the added possibility of on-campus spread, particularly amongst roommates. Based on the weekly screening and diagnostic testing of residential students, we analyzed the transmission of COVID-19 in the residence halls.

### Lower infection rate in single occupancy rooms

We first examined whether students living in single rooms versus those living in multiple occupancy rooms became infected at different rates. Approximately 30% of the 6408 residential students lived in single rooms (1916 students) and 70% in multiple occupancy rooms (4492 students in 2230 rooms). Of the students in single rooms, we detected198 positives (10.3%). By contrast, we detected 860 positives (19.1%) in multiple occupancy rooms (Fig. 1A, Fig. 2, and Table 1). To determine whether this observed difference in infection rate could be due to a testing bias toward students in multiple occupancy rooms, we calculated the fraction of required weekly tests on record for each student. We found that students living in singles tested at a slightly higher frequency than students in multiple occupancy rooms (mean singles = 0.54; mean multiple occupancy = 0.51; Fig. 1B). Test frequency bias is therefore unlikely to be responsible for the higher observed cumulative positivity among students in multiple occupancy rooms.

**Table 1.** Population characteristics.

| Category | Category Description | Number | Percent (%) | Percent Calculation | Percent Description |
| --- | --- | --- | --- | --- | --- |
| A | Total residential students | 6408 | – | – | – |
| B | Total students living in single rooms | 1916 | 29.9 | B/A | Fraction of residential students living in single rooms |
| C | Total students living in multiple occupancy rooms | 4492 | 70.1 | C/A | Fraction of residential students living in multiple occupancy rooms |
| D | Positive residential students | 1058 | 16.5 | D/A | Fraction of residential population with SARS-CoV-2 infection detected |
| E | Positive students in single occupancy rooms | 198 | 10.3 | E/F | Fraction of single occupancy rooms with infected student; fraction of students living in single occupancy rooms who were positive |
| F | Positive students in multiple occupancy rooms | 860 | 19.1 | F/G | Fraction of students living in multiple occupancy rooms who were positive |
| G | Total occupied rooms | 4146 | – | – | – |
| H | Total single occupancy rooms | 1916 | 46.2 | H/G | Fraction of occupied rooms that are singles |
| I | Total multiple occupancy rooms | 2230 | 53.8 | I/G | Fraction of occupied rooms that are multiple occupancy |
| J | Rooms with at least 1 positive student | 875 | 21.1 | J/G | Fraction of occupied rooms with one or more infected students |
| K | Single rooms with a positive student | 198 | 22.6 | K/J | Fraction of rooms with a positive student that were single occupancy |
| L | Multiple occupancy rooms with at least 1 positive student | 677 | 77.4 | L/J | Fraction of rooms with one or more infected students that are occupied by >1 student |
| M | Rooms with multiple positive students | 181 | 26.7 | M/L | Fraction of multiple occupancy rooms with >1 infected student |
| N | Rooms with multiple positive students detected same day | 44 | – | – | – |
| O | Rooms with multiple positive students detected >14 d apart | 21 | – | – | – |
| P | Multiple occupancy rooms with only 1 positive student | 496 | – | – | – |
| Q | Rooms from (R) where presumptive negative roommate has an insufficient testing record | 98 | – | – | – |
| R | Rooms from (R) where presumptive negative roommate has an insufficient testing record | 5 | – | – | – |
| S | Rooms from (P) excluded from analysis (Q + R) | 103 | – | – | – |
| T | Multiple occupancy rooms with at least 1 positive student and a sufficient testing record to assess transmission (L-S) | 574 | – | – | – |
| U | Rooms with multiple positive students likely infected externally and simultaneously (N) | 44 | 7.7 | U/T | Fraction of multiple occupancy rooms with >1 infected student that do not likely represent in-room transmission |
| V | Rooms with potential in-room transmission (M - N - O) | 116 | 20.2 | V/T | Fraction of multiple occupancy rooms with >1 infected student that likely transmitted in the room |
| W | Rooms with sufficient testing record with unlikely in-room transmission (P - Q), 1 positive student | 398 | – | – | – |
| X | Rooms with multiple positive students detected >14 d and with likely co-habitation during index case's infectious period (O - R) | 16 | – | – | – |
| Y | Rooms with sufficient testing record with unlikely in-room transmission (W + X) | 414 | 72.1 | Y/T | Fraction of multiple occupancy rooms with 1 infected student detected and sufficient testing record of roommate pair; unlikely in-room transmission |
|  | Correcting for rooms with insufficient testing record |  |  |  |  |

**Fig. 1.**
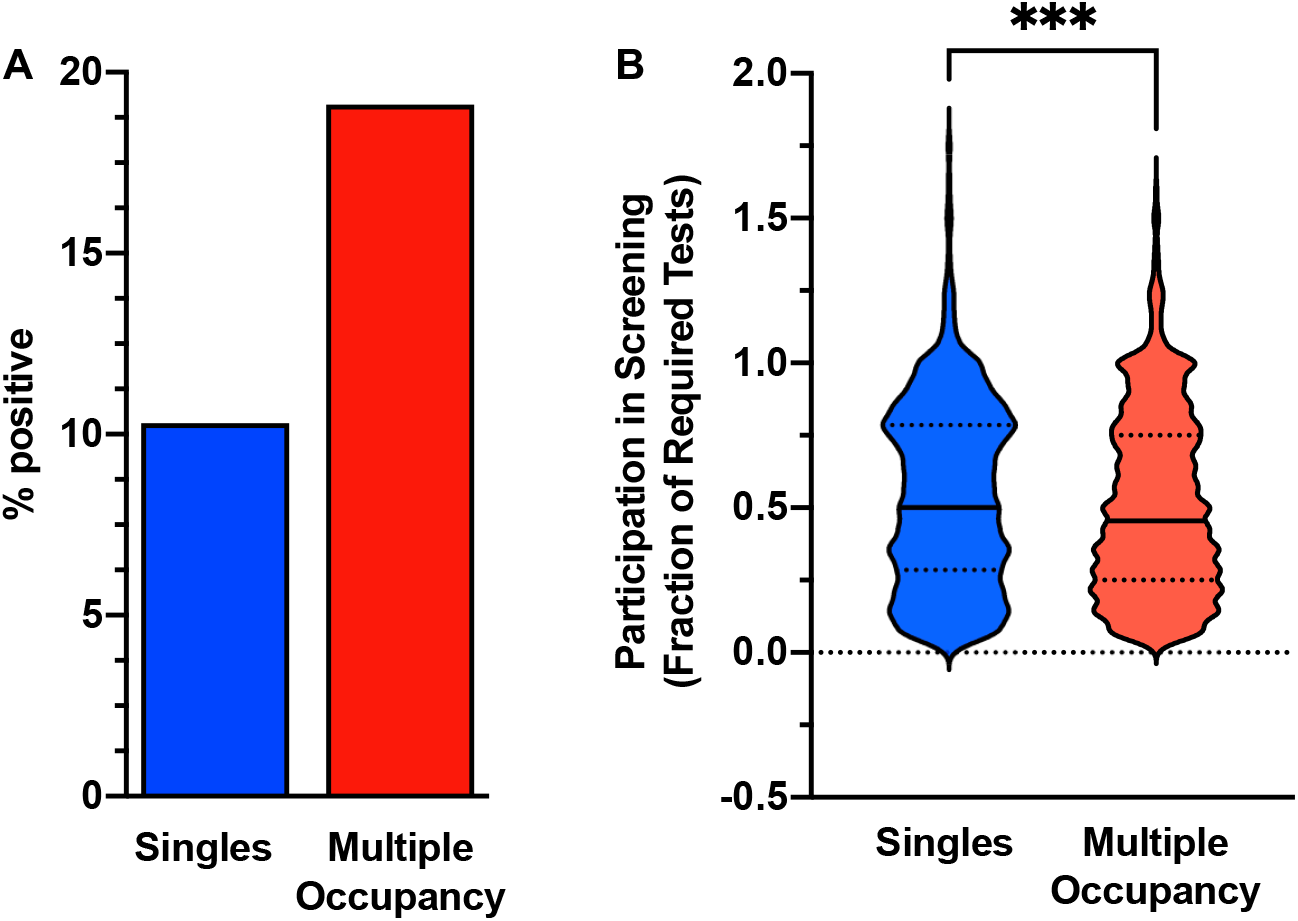
Students in multiple occupancy rooms were infected at twice the rate of students in single rooms. **(A)** 198 students in 1916 single occupancy rooms (10.3%) were infected. 860 students in multiple-occupancy rooms were infected out of a total of 4492 students (19.1%). **(B)** The mean and median fraction of required tests were modestly higher for students in single compared to multiple occupancy rooms. Violin plots reveal medians (solid lines) and interquartile ranges (dotted lines). Asterisks denote p < 0.001 by unpaired t-test comparing means. n = 1872 for single rooms; n = 4353 for students in multiple occupancy rooms. A small set of students did not have a screening test on record or tested much more frequently than required.

**Fig. 2.**
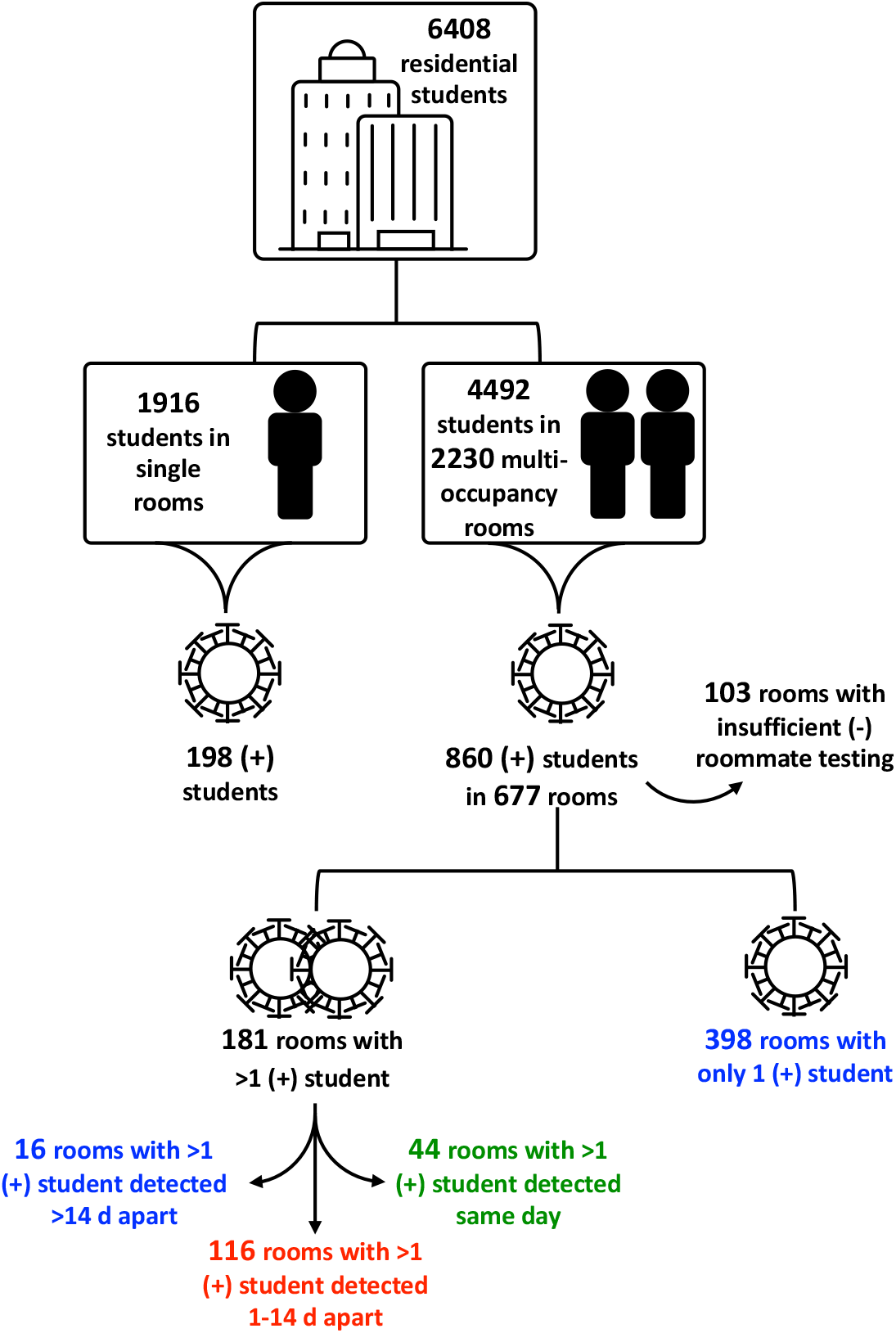
Determining infection patterns in residence hall students. Illustration of logic for identifying rooms with and without potential in-room transmission. (Blue = unlikely in-room transmission; Red = likely in-room transmission; Green = likely simultaneous infection from external source)

### Low rates of roommate transmission

To examine whether inter-roommate transmission accounted for the higher cumulative incidence among students living in multiple occupancy rooms, we investigated the extent to which the timing of cases supported inter-roommate transmission (Fig. 2 and Table 1). The 860 positive students lived in 677 multiple occupancy rooms, of which only 181 rooms had more than one positive student and therefore could potentially represent instances of inter-roommate spread. Of these, we excluded 44 rooms where roommate pairs were detected on the same day as likely same-source external infections, a scenario supported by contact tracing efforts from the CU Pandemic Response Office (manuscript in preparation). We further excluded 21 rooms in which roommates were detected more than 14 days apart as likely independent external infections based on what is known regarding viral incubation time, which also informs the CDC-recommended quarantine time post-exposure (CDC, 2021). Although it is possible that the roommate who became infected later was not actually present in the room during the first-detected roommate’s infectious period, we found a closely-spaced negative test (within the following 2 weeks) for the later roommate in 16 of the 21 rooms. Due to uncertainty in co-habitation during the index case’s infectious period, we excluded the other 5 rooms from further analysis. Therefore, it is likely that these 16 roommate pairs were present together without inter-roommate transmission, and thus that they each were infected by independent sources. Discounting these 65 rooms, we concluded that 116 rooms had potential in-room transmission.

496 multiple-occupancy rooms each had only 1 positive student. To ensure that lack of positive result for the other roommate was not due either to absence from the room during the infectious period or to non-compliance with testing requirements, we excluded 98 rooms where there was not a negative test on record within 3 weeks of the positive roommate’s first date of detection, accounting for incubation period and the weekly RT-qPCR screening cadence. The remaining 398 rooms combined with the 16 rooms with 2 widely-spaced infections represent 414 cases where there was no apparent spread between roommates despite living in close quarters. Overall, of 574 multiple occupancy rooms with at least one positive student and a sufficient testing record, 7.7% had multiple positive cases detected simultaneously and likely due to external exposures of both roommates, 20.2% had likely in-room transmission, and 72.1% did not appear to have transmission events despite close contact during the infectious period (Fig. 3). Thus, the fact that positive cases were nearly twice as likely among students in multiple occupancy rooms compared to singles (Fig. 1) is not primarily due to roommate-to-roommate transmission.

**Fig. 3.**
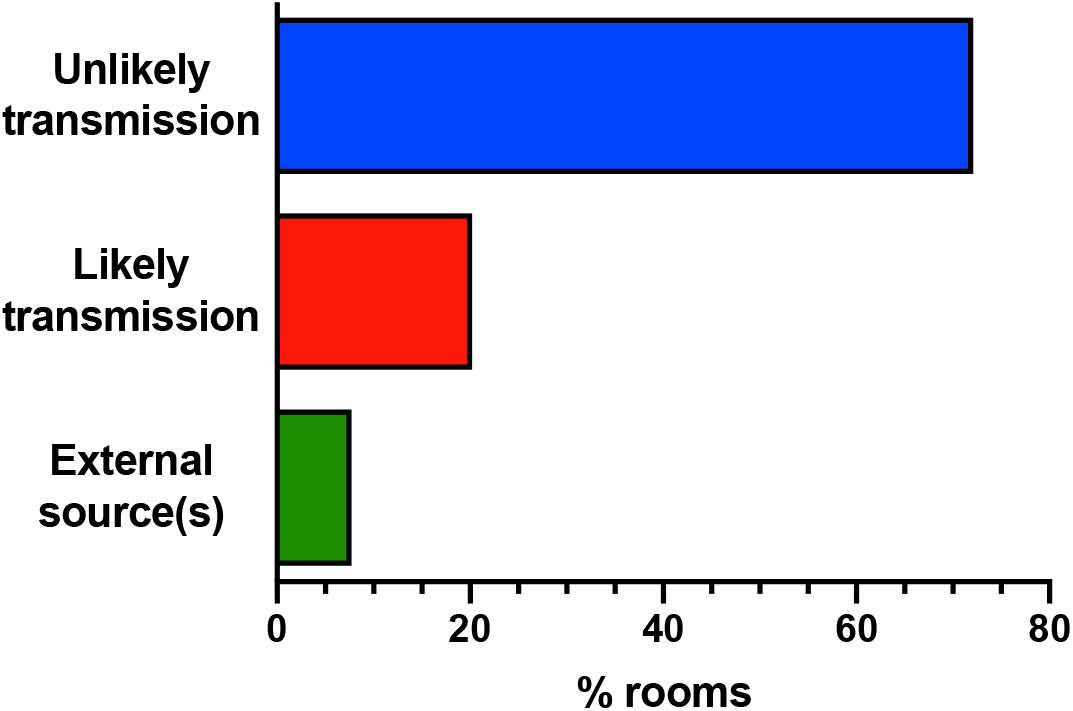
Most multiple occupancy rooms with an infected student did not have inter-roommate transmission. 574 multiple occupancy rooms housed students with a sufficient testing record to evaluate potential transmission. 44 rooms (7.7%) had multiple positive students who were likely infected simultaneously by external sources. 116 rooms (20.2%) had likely in-room transmission from one roommate to another as they were detected within a two-week period but not on the same day. 398 rooms had only one infected student and a closely spaced downstream negative test(s) on record for the roommate, and 16 rooms had 2 widely-spaced infections with clear presence of the second student in the room during the index case’s infections period (72.1% unlikely transmission).

We explored whether the 20% transmission figure could be higher or lower based on analyses that that considered: (1) the possibility that the rooms with same-day detection actually did represent transmission events, (2) the likelihood that two infections from two independent exposures appeared in one room by random chance, and (3) the potential that a mid-semester remote learning period could have prevented some transmission events (see Supplemental Information for more detailed description). Taken together, our analysis argues that transmission of COVID-19 in asymptomatic college students is a low probability event with only between 20 and 28% of the cases of one infected roommate actually transmitting disease to their co-occupant.

### Higher viral load correlates with higher transmission risk

One factor that could have contributed to a secondary attack rate of only 20%-28% is diversity in viral load. Multiple reports have indicated that viral load can differ by orders of magnitude across patients (Kissler et al., 2020; Lennon et al., 2020). Moreover, there is growing evidence amongst symptomatic patients that higher viral load can result in a higher risk of transmission (Kawasuji et al., 2020; Marks et al., 2021). To compare viral load in these students, we looked at the quantification of viral E gene RNA in the screening RT-qPCR test as a proxy for viral genomes. We chose the lowest C_q_ on record for each room in both the likely and unlikely transmission groups. C_q_ values indicate how many amplification rounds were necessary to reach detection, so higher C_q_ values indicate lower viral loads and each whole unit is a factor of 2 in RNA copies per ml. As shown in Fig. 4, we found that the average viral load was 6.5-fold higher in rooms with likely transmission (mean C_q_ = 26.2) than in rooms without transmission (mean C_q_ = 28.9). The difference in median [IQR] between groups was similar (likely transmission = 26.11 [23.36 – 29.13]; unlikely transmission = 29.32 [26.34 – 31.64]). This striking difference between groups indicates that individuals with higher viral load may indeed result in a higher effective dose for people exposed to them and raise their risk of infection accordingly.

**Fig. 4.**
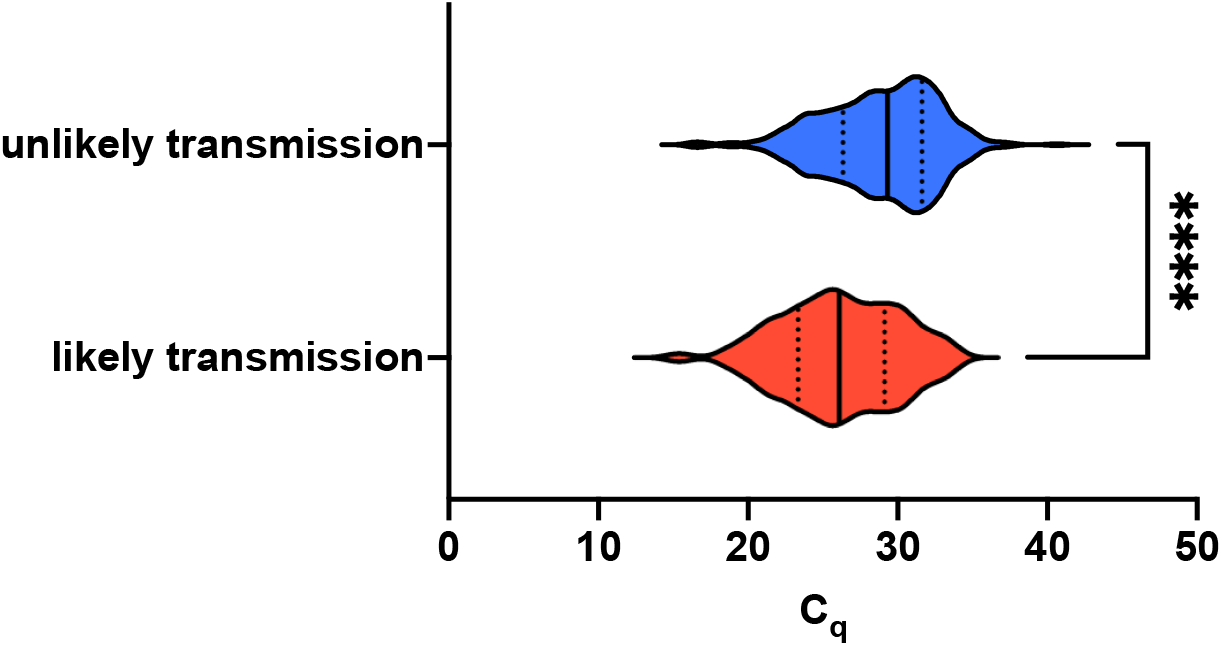
Rooms with transmission had higher viral load. Comparison of the lowest C_q_ on record in multiple occupancy rooms with or without likely transmission for students detected through the screening program. The significant difference between groups indicated a 6.5-fold higher average viral load in the group with transmission. Violin plots reveal medians (solid lines) and interquartile ranges (dotted lines). Asterisks denote p < 0.0001 by unpaired t-test comparing means. n = 80 for likely transmission; n = 366 for unlikely transmission.

These cases also spanned a range of over 7 orders of magnitude in viral load, with the highest load found in the likely transmission group (C_q_ = 15.4) and the lowest found in the unlikely transmission group (C_q_ = 40.6). The only cases with C_q_ greater than 34 (22 cases) were found in the unlikely transmission group.

The correlation with viral loads is striking given that our weekly testing cadence typically allows for only one viral load measurement for each infected individual. These measurements could occur at different times over the course of infection, during which viral loads may vary greatly from one day to the next (Kissler et al., 2020). However, because our sample set comprises hundreds of identified infections, C_q_ values should be representative of the window of an infection during which individuals can test positive. Moreover, due to the regular screening program combined with the high sensitivity of the assay (5 virions/µL), it is likely that most students were in the early stages of infection when detected. We found that 48% of students with unlikely transmission and 56% of students with likely transmission had a negative test on record from the previous week, and these numbers increase to 81% and 78%, respectively, when considering a two-week window. There was no significant difference in the distributions of most recent negative test dates between the two groups (two sample Kolmogorov-Smirnov test; p = 0.62, D statistic = 0.093). As a result, the observed higher viral loads among transmitting roommate pairs are unlikely to be an artifact of biases in sampling times.

### Time to isolation is not correlated with increased transmission

Increased time of exposure to an infected person is one of the factors used to identify at-risk close contacts and invoke quarantine protocols (CDC, 2021). CU Boulder established dedicated isolation residence halls to temporarily re-house infected students in an effort to reduce community spread, particularly amongst roommates. Isolation was not initiated until infected students were confirmed by a diagnostic RT-qPCR test, which led to variation in the time before a student left their home residence hall and went into isolation. Therefore, we examined whether there was any difference between groups in the screening testing or directly through diagnostic testing) and the date that individual entered isolation.

We found no difference in time-to-isolation between rooms with and without transmission (permutation hypothesis test, p = 0.19; see Methods). Furthermore, although 18.6% of unlikely transmission cases entered isolation on the day they were detected, compared to only 9.6% of the transmission cases, a Kaplan-Meier analysis of the time-to-isolation distributions indicated no significant difference between the likely and unlikely transmission groups (Fig. 5; Gehan-Breslow-Wilcoxon test, p=0.26). While this does not negate the potential benefit of isolating infected individuals from their households, it does indicate that a lag in time-to-isolation does not explain the transmissions we observed.

**Fig. 5.**
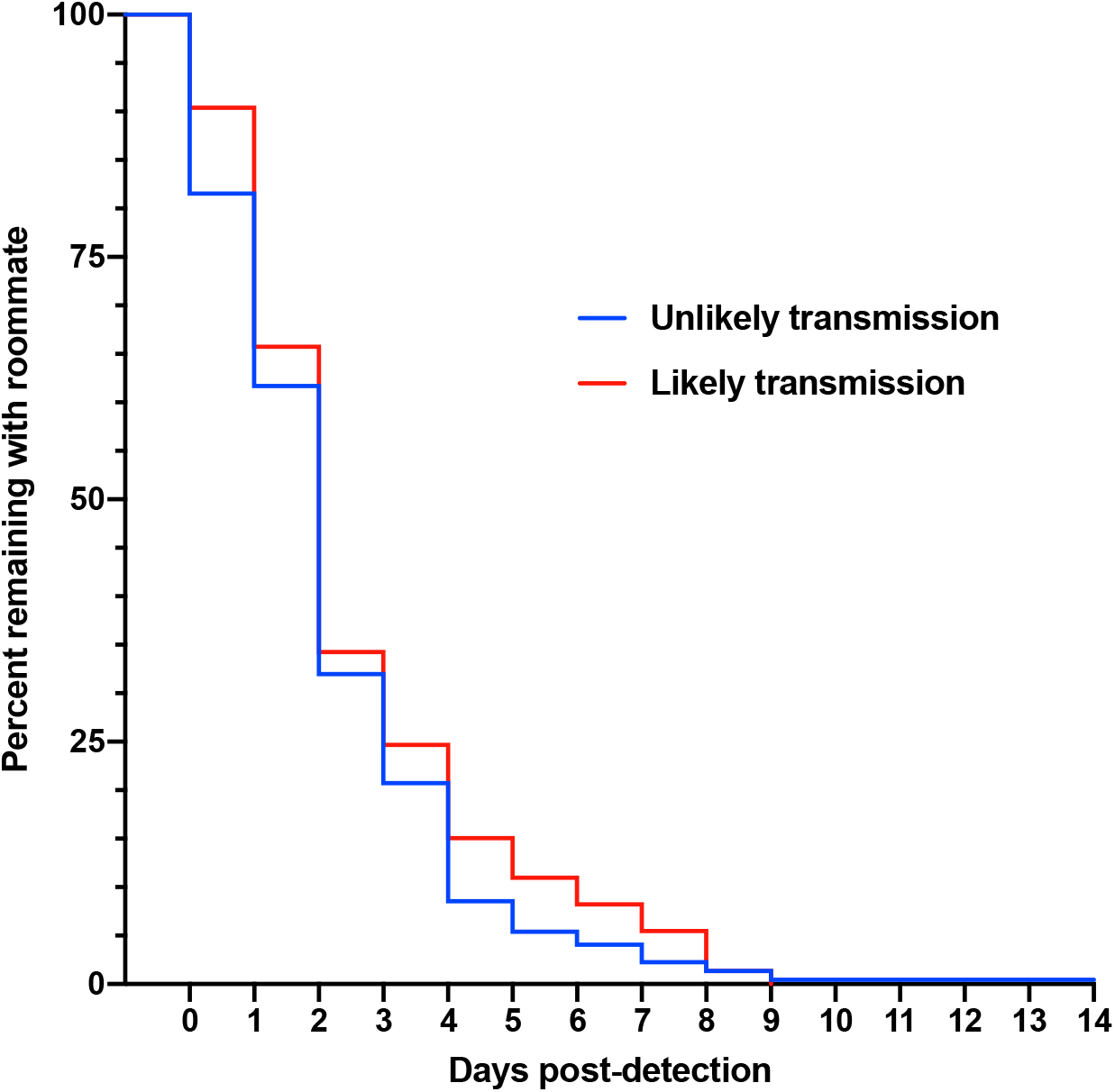
There was no difference in isolation patterns between rooms with and without transmission. Kaplan-Meier curves comparing the percent of positive cases remaining with their roommate versus the days since first detection. There is no significant difference between curves (p = 0.26 by Gehan-Breslow-Wilcoxon test). n = 73 for likely transmission; n = 222 for unlikely transmission.

## DISCUSSION

This study revealed several key findings that help to identify which factors influence transmission from largely asymptomatic cases living in small, dense households. First, we observed a 2-fold higher positivity amongst students with roommates than those who live alone. However, this difference cannot be explained solely by transmission between roommates, since we estimate such transmission only occurred in 20% of these multiple occupancy rooms. Interestingly, behavioral studies have shown that people who live alone have less frequent social contact than people who live in households and that roommates can exert peer effects with risky behaviors (Eisenberg et al., 2014; Hefner and Eisenberg, 2009). Therefore, it may be that students living with roommates are at a higher risk of infection due to both higher levels of social contact outside the home and moderate levels of inter-roommate transmission within the home.

Second, a notable contribution of this work is to provide evidence that viral levels are positively correlated with the probability of transmission in the context of a high frequency screening program. There has been very little work examining the relationship between viral load and the risk of transmission, although it is a common hypothesis underpinning COVID-19 transmission modeling studies (Larremore et al., 2021, 2020). We observed that students who transmitted SARS-CoV-2 to their roommates had, on average, 6.5-fold higher viral loads than students who did not transmit virus to their roommates (Figure 4). Thus, we suggest that a key parameter in transmission between roommates is the viral load of the index case in each room. We note that this time period precedes widespread U.S. circulation of mutant SARS-CoV-2 variants that arose in the U.K. (strain B.1.1.7), South Africa (B.1.351), and Brazil (P.1). These variants of concern could have different transmission dynamics and virulence.

The importance of viral load in COVID-19 transmission is beginning to be supported by other recent analyses. A recent study of a small group of 21 adults found higher viral load at initial sample collection in the 14 people who transmitted infection compared to those who did not (Kawasuji et al., 2020). However, these people were initially identified from hospitalized patients and subsequent contact tracing, so there is some uncertainty with respect to timing of collection relative to stage of infection. Another study of 282 mildly symptomatic, but not hospitalized, patients found that viral loads greater than 1×10^10^ copies/mL were associated with double the rate of transmission (24%) than viral loads below 1×10^6^ copies/mL (Marks et al., 2021). Our study supports these observations by bringing new evidence from 579 concordant and discordant sets of asymptomatic young adults undergoing regular RT-qPCR testing.

It is also worth highlighting that our study population was largely asymptomatic, which is a continued concern as a COVID-19 transmission source and yet is relatively understudied compared to symptomatic, often hospitalized, patients. For example, in a recent meta-analysis of 79 studies, 73 focused on hospitalized patients (Cevik et al., 2021). We were able to identify asymptomatic infected individuals due to a mandatory weekly screening program for all residential students and robust contact tracing, both coupled to on-campus diagnostic testing.

The majority of students were asymptomatic at the time of first detection. We found transmission rates similar to studies involving mostly symptomatic populations, supporting the idea that asymptomatic infected individuals can transmit as efficiently as symptomatic individuals (Madewell et al., 2020). This is strengthened by the observation that viral loads are similar between symptomatic and asymptomatic individuals (Lennon et al., 2020; Yang et al., 2021).

The large fraction of rooms that did not experience transmission is encouraging as roommate pairing could support positive mental health, particularly with restricted in-person socialization opportunities. In fact, when the Georgia Institute of Technology offered its students the opportunity to leave their roommates and move into single rooms after they detected a wave of cases, many elected to stay co-housed for mental health reasons, despite a 30% secondary attack rate amongst roommates (Gibson et al., 2021). Georgia Tech has a residential student population comparable to CU Boulder. Notably, the higher inter-roommate transmission seen at Georgia Tech (∼30%) as compared to CU Boulder (∼20%) occurred with a lower cumulative positive rate at Georgia Tech (9.7% amongst students) than observed at CU Boulder (16.5%). However, participation in weekly screening was mandatory at CU Boulder but not at Georgia Tech. Thus, additional cases might have been missed at Georgia Tech.

In contrast to the importance of viral load, we did not observe a significant difference in isolation speed between the group that experienced transmission and the group that did not. This does not mean that isolation has no effect on preventing transmission. By comparison to the 20% secondary attack rate we saw among roommates, the secondary attack rate within households is reported to be 38% between spouses but 18% between non-spouse household members (Madewell et al., 2020). The reasons for this difference are unknown, but spouses likely spend more time in close contact with each other, including sleeping in the same room. Because roommates in residence halls also share sleeping quarters, this suggests that transmission could have been higher in the absence of off-site isolation.

Given the many unknowns surrounding an emergent disease, concerns regarding the safety of roommate pairing were justifiable early in the pandemic as universities made operational decisions in anticipation of Fall 2020 terms. However, there are multiple advantages to sharing a residence hall room. Many students experience mental health benefits from this socialization opportunity, particularly in the background of gathering restrictions that were incorporated into community and university public health measures. In addition, allowing roommate pairing permits higher density on-campus presence, which is valuable for students who benefit from in-person instruction, added university support services, or who may experience housing, food, or financial insecurities at their permanent residence. Our study adds reassurance that roommate pairing is relatively safe, with surprisingly low rates of inter-roommate SARS-CoV-2 transmission. This is qualified by the finding that asymptomatic individuals with high viral load do pose a transmission risk. Therefore, quantitative tests than can efficiently uncover these cases can have great value in prioritizing downstream measures such as isolation protocols, contact tracing, and quarantine advice for close contacts, particularly as it is estimated that 80% of secondary transmission events originate from ∼10% of infected individuals (Endo et al., 2020). Those with highest viral load could represent potential super-spreaders, and efficiently identifying them could help prevent large outbreaks.

## MATERIALS AND METHODS

### CU Boulder Public Health Operations

#### RT-qPCR Screening

Residential students were required to test weekly with each residence hall assigned to a day of the week, thus treating each hall as a cohort. The screening test was developed at CU Boulder and is described in detail elsewhere, including primer and probe sequences and fluorophores along with example standard curves (Yang et al., 2021). Briefly, students self-collected 1 mL saliva under observation at designated on-campus sites. Raw saliva was heat-inactivated on-site for 30’ at 95°C and subsequently stored on ice. Saliva was mixed 1:1 with 2X TBE (Tris-borate-EDTA) and 1% tween-20 for final concentrations of 0.5X saliva/1X TBE/0.5% tween-20. The CU-Saliva PCR Integrated-Triplex Test was used to measure two SARS-CoV-2 viral RNA targets (envelope (E) and nucleocapsid (N1 or N2, depending on supply chain availability)) and one human control (RNaseP). Primers were taken from FDA-approved EUAs. Reverse transcription combined with Taqman-based qPCR was employed for triplex assays (TaqPath™ 1-Step Multiplex Master Mix (No ROX); Fisher Scientific, A28523) run on a BioRad CFX96 or CFX384 Real Time PCR Detection System. C_q_ values were determined with the CFX Maestro’s built-in Regression C_q_ Determination Mode. This assay has a 5 virions/µL limit of detection. Presumptive positive calls were made if both viral targets and RNaseP were detected while “no signs detected” calls were made if neither viral target amplified but RNaseP did. Presumptive positive students were referred to medical staff at the CU Public Health Clinic for contacting and scheduling follow-up diagnostic testing.

#### Diagnostic RT-qPCR Testing

Diagnostic RT-qPCR testing was available at the on-campus Public Health Clinic through Student Health Services. The Lyra Direct SARS-CoV-2 Assay (Quidel; M124) was used according to the EUA, including guidance on positive/negative calls. Students self-collected nasal specimens by swabbing the anterior nares under observation in the clinic. The assay measures one viral target (pp1ab) and one control target (MS2 bacteriophage). It was run on a ThermoFisher QuantStudio 7 Pro. The test audience included presumptive positive students referred from screening, symptomatic students, and students identified through contact tracing.

#### Isolation Protocol

Students diagnosed as SARS-CoV-2 positive with the Lyra Direct assay were moved into isolation accommodations for 10 days in accordance with CDC recommendations (CDC, 2021). They either isolated in dedicated residence halls with single rooms containing en suite bathrooms or were allowed to temporarily move home if they lived within a 250-mile radius.

#### Data Collection

SARS-CoV-2 test data, housing assignments, and isolation data were collected through the campus operational framework and were all stripped of identifying information before proceeding with data analysis. Data were analyzed and reported here in aggregate.

#### Data Analysis

##### Fraction of Required Tests

Students were present for 14 weeks on campus and required to test weekly as long as they continued to test negative. If students tested positive through a diagnostic test, they were exempt from testing for the rest of the semester as that was within the 90-day recommended test abstention window based on viral RNA persistence during protracted viral clearance periods. For every residential student, we determined the number of required tests adjusted to their detection date if they did test positive. We then determined the total number of tests on record for each student and calculated the fraction of required tests that value represented. A small number of students chose to test much more frequently than once a week, so we performed an outlier analysis with the ROUT method in GraphPad Prism, which is based on false discovery rate, in order to ensure that they did not artificially skew the group means and medians (Motulsky and Brown, 2006). We removed 17 high frequency testers from the 1889 students in single rooms and 96 from the 4449 students in multiple occupancy rooms. We assessed statistical significance of differences in test compliance between the remaining 1872 students in single rooms and the 4353 students in multiple occupancy rooms by unpaired t-test and further compared the groups with a violin plot, displaying medians and interquartile ranges.

##### In-Room Transmission

For positive residential students we determined the first date of detection by compiling all positive tests on record from both screening and diagnostic testing. We used this date to calculate the detection interval between positive roommate pairs and the minimum amount of time spent in a room while positive before moving to isolation.

To determine transmission likelihood, we defined likely transmission as two positive roommates detected within a 1 to 14-day range, based on viral latency, detection thresholds, and the weekly screening cadence. If two positive roommates were detected on the same day, we considered them as having been infected by the same external source rather than inter-roommate transmission. If two positive roommates were detected more than two weeks apart, we considered them as having been infected by two independent, external sources rather than inter-roommate transmission. We also looked for presence of the later positive in the testing record around the time of detection of the index case as a surrogate measure of on-campus presence. If a multiple occupancy room had only one positive roommate, we considered them cases of unlikely transmission as long as the negative roommate was present in the testing record within the three weeks following detection of the positive roommate. This was done to minimize uncertainty of their presence in the room during the positive roommate’s infectious period.

To calculate the expected number of multiple occupancy rooms with 2 students infected by chance through external exposures, we first determined the campus infection rate (*p*) for each week (*t*) of the semester. Then we compiled the number of multiple occupancy rooms with no infected students remaining each week (*n*), as this represents the actual number of rooms at risk each week. The probability of 2 students becoming infected from independent events is *p*^*2*^, and multiplying that by *n* yields the number of rooms expected per week. The sum of these weekly numbers results in the total rooms (*N*) predicted to have 2 independent infection events over the 14 weeks of the semester. This is summarized by the following formula:

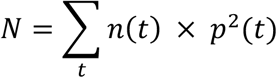

##### Time to Isolation

To determine time to isolation, we calculated the difference between the first date of detection in a room and the date that student entered isolation. Isolation data were not available for every student. We calculated this number for the likely transmission and the unlikely transmission groups. We analyzed the time distributions of these two groups by permutation hypothesis testing in R Studio, permuting the random dataset 100,000 times. We calculated the p-value based on the difference in means in the experimental dataset compared to the permutation dataset. We also analyzed the isolation kinetics with a Kaplan-Meier plot followed by a Gehan-Breslow-Wilcoxon test to compare the curves.

##### Viral Load

We had quantitative data available from the screening test, so we analyzed the lowest C_q_ recorded in each multiple occupancy room as a surrogate for the highest viral load. We compared the mean C_q_ in the likely transmission to the unlikely transmission group. We assessed statistical significance of the mean C_q_ difference with an unpaired t-test and further compared the groups with a violin plot, displaying medians and interquartile ranges.

##### Software

We used Bio-Rad CFX Maestro 2.0 (5.0.021.0616) for Windows to collect RT-qPCR screening results. We performed all graphing and statistical analysis, except for permutation hypothesis testing, with GraphPad Prism version 9.0.1 (128) for macOS (graph and test types identified in the relevant Methods section, Results section, and figure legends). We performed permutation hypothesis testing in R Studio Version 1.4.1103 for macOS using R 4.0.3 GUI 1.73 Catalina build (7892).

## Data Availability

Data are available upon request.

## SYMBOLS AND ABBREVIATIONS

C_q_ =: quantification cycle
CDC =: Centers for Disease Control
COVID-19 =: Coronavirus Disease-19
CU =: University of Colorado
E =: envelope
FDA =: Food and Drug Administration
IQR =: interquartile range
N =: nucleocapsid
pp1ab =: replicase polyprotein 1ab
RT-qPCR =: Reverse transcription quantitative polymerase chain reaction
SARS-CoV-2 =: Severe Acute Respiratory Syndrome Coronavirus 2

## ACKNOWLEDGEMENTS

We are grateful to Carolyn Decker for developing the reliable and sensitive RT-qPCR screening test upon which the screening program at CU Boulder was built. We also thank CU leadership, the Scientific Steering Committee, and the Pandemic Response Office for institutional support and scientific guidance that allowed the university to remain operational. Finally, we acknowledge the CU Boulder student body for persevering through an unprecedented college semester and participating in the CU public health system.

We dedicate this work to the memory of Denise Muhlrad, who was essential in establishing the screening program despite the many challenges a pandemic presents.

## ETHICS STATEMENT

Data on student participants were aggregated from University of Colorado Boulder operational COVID-19 screening activities. This does not meet the definition of human subject research described in the United States Health and Human Services 45 Code of Federal Regulations Part 46. For this reason, this study did not fall under Institutional Review Board purview.

## COMPETING INTERESTS

The following authors have financial ties to companies offering commercial SARS-CoV-2 testing: TKS, EL, and PKS are co-founders of TUMI Genomics. RP is a co-founder of Faze Medicines.

## FUNDING

This work was supported by US CARES Act funding to CU Boulder.

## Notes

### Author Declarations

Claire Dunne, Ph.D. IRB Program Director University of Colorado Boulder http://www.colorado.edu/researchinnovation/irb The project you describe would be regarded as part of campus operations and not research as defined at 45CFR46.102(l) as it was not designed to develop or contribute to generalizable knowledge. https://www.hhs.gov/ohrp/sites/default/files/revised-common-rule-reg-text-unofficial-2018-requirements.pdf For your reference I have included the definitions at 45CFR46.102(l) Research means a systematic investigation, including research development, testing, and evaluation, designed to develop or contribute to generalizable knowledge. Activities that meet this definition constitute research for purposes of this policy, whether or not they are conducted or supported under a program that is considered research for other purposes. For example, some demonstration and service programs may include research activities. For purposes of this part, the following activities are deemed not to be research: (1) Scholarly and journalistic activities (e.g., oral history, journalism, biography, literary criticism, legal research, and historical scholarship), including the collection and use of information, that focus directly on the specific individuals about whom the information is collected. (2) Public health surveillance activities, including the collection and testing of information or biospecimens, conducted, supported, requested, ordered, required, or authorized by a public health authority. Such activities are limited to those necessary to allow a public health authority to identify, monitor, assess, or investigate potential public health signals, onsets of disease outbreaks, or conditions of public health importance (including trends, signals, risk factors, patterns in diseases, or increases in injuries from using consumer products). Such activities include those associated with providing timely situational awareness and priority setting during the course of an event or crisis that threatens public health (including natural or man-made disasters).

