## Supplemental Information for "Higher viral load drives infrequent SARS-CoV-2 transmission between asymptomatic residence hall roommates"

**Sensitivity Analysis**

If we exclude the 44 rooms with same-day detection as likely same-source external infection, there are 530 rooms with the potential for transmission. In this case, 21.9% (116) of the rooms experienced inter-roommate spread, and 78.1% (414) of multiple occupancy rooms with at least one infected student did not likely result in inter-roommate spread. In the most conservative case, if the 44 rooms with same-day detection actually do represent inter-roommate spread, then 160 of 574 rooms (27.9%) experienced spread and 72.1% did not.

Two additional observations suggest that these 116 cases in the same room involve inter-roommate transmission. First, we can predict the expected number of multiple occupancy rooms where two roommates become infected independently by external exposures. Based on the weekly infection incidence and the weekly number of rooms at risk of infection, we estimate that 14 rooms would have had both roommates infected through independent events over the course of the semester (see Materials and Methods for a full description of the calculation). This is much lower than the observed 116 rooms with likely inter-roommate transmission, and is similar to the 16 rooms with widely spaced infections that likely resulted from two separate external exposures. Second, the events we considered as inter-roommate transmission occurred within 1-14 days of each other, consistent with inter-roommate transmission given the kinetics of viral infectivity. Thus, while it is possible that some of our 116 events reflect two external infection events, their prevalence and timing argue that these are predominantly intra-room transmission events.

One caveat to our analysis is we do not have direct knowledge of students living in their assigned residence hall rooms. However, because we considered only those students with an ongoing residence hall screening record, we are certain they were present in the local community and most likely living in their assigned residence hall room. Moreover, we can estimate the worst-case scenario for students being absent from their residence hall rooms since the university monitored the number of Wi-Fi connections from the residence halls over the course of the semester. We observed that residence hall density was reduced by 33% during a period of the semester when the university temporarily shifted to online-only classes. 80 of the rooms with unlikely transmission (defined as either a positive/negative roommate pair or roommate pairs who became positive at a widely spaced interval) were detected during this remote learning period. Thus, at a lower limit perhaps 33% of the cases in these rooms might have been prevented due to a roommate absence. In this more conservative calculation, 24.9% of the rooms (143/574) would have had intra-room transmission during the semester and 67.4% would have not (387/574).
